# Epigenome-wide association study of serum folate in maternal peripheral blood leukocytes

**DOI:** 10.1101/2022.11.23.22282639

**Authors:** N. Fragoso-Bargas, C.M. Page, B.R. Joubert, S.J. London, S. Lee-Ødegård, J.O. Opsahl, L. Sletner, A.K. Jenum, E. Qvigstad, R.B. Prasad, G.-H. Moen, K.I. Birkeland, C. Sommer

**Author notes:** Corresponding author: Christine Sommer, PhD, Department of Endocrinology, Morbid Obesity and Preventive Medicine, Oslo University Hospital, Postbox 4959 Nydalen, N-0424 Oslo, Norway.

## Abstract

**Aim:** To perform an epigenome-wide association study (EWAS) of serum folate in maternal blood.

**Methods:** We performed cross-ancestry (Europeans=302, South Asians=161) and ancestry-specific EWAS in the EPIPREG cohort, followed by methyl quantitative trait loci (mQTL) analysis and association with cardiometabolic phenotypes. We attempted replication using folate intake estimated from a food frequency questionnaire and maternal blood methylation data from MoBa, and in a previous published EWAS of maternal serum folate in cord blood.

**Results:** cg19888088 (cross-ancestry) in *EBF3*, cg01952260 (Europeans), and cg07077240 (South Asians) in *HERC3* were associated with serum folate. cg19888088 and cg01952260 were associated with diastolic blood pressure. cg07077240 was associated with variants in *CASC15*. The findings were not replicated in the independent samples.

**Conclusion:** Serum folate was associated with methylation at three CpG sites.

## Introduction

Folate (vitamin B9) is a coenzyme in the one-carbon metabolism, a pathway that provides one-carbon units for nucleotide biosynthesis and methylation reactions [1,2]. The methyl groups derived from folate are necessary to form SAM (S-Adenosyl methionine) which is a universal carbon donor for DNA methylation [3]. DNA methylation is an epigenetic mechanism that regulates gene expression and genome stability, as it represses repetitive elements [4]. Adequate folate intake helps to maintain normal DNA methylation levels and minimise DNA damage [5].

It is well recognized that folate is important for tissue growth and fetal development during pregnancy [6]. However, previous studies suggest that folic acid supplementation is also associated with decreased fasting glucose, fasting insulin and insulin resistance[7], and reduced risk for stroke and cardiovascular disease [8]. It has been proposed that DNA methylation could be an important mechanism underlying the observed associations between folate and cardiometabolic traits [9,10]. In line with this hypothesis, a study suggested that low folate levels were associated with hypo methylation in several CpG sites in liver cells of patients with type 2 diabetes [11].

To date, there is only one epigenome-wide association study (EWAS) of serum folate, which examined the association between maternal serum folate levels and differential DNA methylation in cord blood of their offspring [12]. Another study performed in cord blood, evaluated the association between folate supplementation and DNA methylation during pregnancy [13]. However, there is no EWAS of serum folate performed in peripheral blood leukocytes.

The epigenomic background of serum folate levels could give valuable insights into folate regulation. Hence, the aims of this study were to 1) perform an EWAS of serum folate in maternal peripheral blood leukocytes, 2) evaluate if the methylation of CpG sites associated with folate were related to cardiometabolic phenotypes, 3) elucidate if methylation of the CpG sites were genetically regulated, 4) evaluate if our findings are replicated in maternal peripheral blood leukocytes with maternal estimated folate intake data assessed with FFQ as well as in cord blood with maternal serum folate data.

## Methods

### Study Population

The STORK Groruddalen (STORK G) study is a population-based cohort which included 823 healthy pregnant women in the multi-ethnic area of Groruddalen in Oslo, Norway, from 2008-2010. STORK G has been described in detail in previous papers [14]. The inclusion criteria were that the woman: 1) lived in the study district; 2) planned to give birth at one of the two study hospitals; 3) were less than 20 weeks pregnant; 4) could communicate in Norwegian or any of the eight translated languages; 5) were able to give informed consent. Women were enrolled during early pregnancy, and women with pre-gestational diabetes, or in need of intensive hospital follow-up during their pregnancy were excluded. Self-reported ethnic origin was defined by either the individual’s country of birth or their mother’s country of birth if the latter was born outside Europe.

The Epigenetics in Pregnancy (EPIPREG) sample included all women of European (n=312) or South Asian (n=168) ancestries participating in STORK G with available DNA [15].

STORK G, including the genetic, and epigenetic sub-studies (EPIPREG), was approved by the Norwegian Regional Committee for Medical Health Research Ethics South East (ref. no. 2015/1035). Written informed consent was given by all participants.

### Folate measurement

Folate was measured in biobanked serum collected at gestational week 28 with electrochemiluminescence (ECLIA) (Roche diagnostics international at Medical Biochemistry, Oslo University Hospital. Folate deficiency was determined at <7 nmol/L [16].

### Cardiometabolic phenotypes

The data of the cardiometabolic phenotypes used in this study were collected and measured in gestational week 28. Venous blood was drawn into ethylenediaminetetraacetic acid (EDTA) tubes. Then, the samples were either biobanked or subject to further preparation and analyses. All women underwent a 75g oral glucose tolerance test. GDM was diagnosed with the WHO 1999 criteria (fasting glucose ≥ 7.0 mmol/l and/or 2-hour glucose ≥ 7.8 mmol/l). Fasting insulin was measured with non-competing immunofluorometric assays (DELFIA, PerkinElmer Life Sciences, Wallac Oy, Turku, Finland). Insulin resistance was estimated by the homeostasis model of insulin resistance, using the HOMA2 calculator version 2.2.2 (https://www.dtu.ox.ac.uk/homacalculator) based on fasting glucose (HemoCue, Angelholm, Sweden) and C-peptide (DELFIA, PerkinElmer Life Sciences, Wallac Oy, Turku, Finland) [17]. Fasting plasma total cholesterol, HDL cholesterol and triglycerides levels were measured with a colorimetric method (Vitros 5.1 fs, Ortho clinical diagnostics, Neckargemünd, Germany) at Akershus University Hospital, Lørenskog, Norway. LDL cholesterol was calculated with Friedewald’s formula [18,19].

Anthropometric data in the present study were also measured at gestational week 28. Systolic and diastolic blood pressure were measured with M6 Comfort HEM-7000-E (Omron, Kyoto, Japan). Body mass index (BMI) was calculated from measured height using a fixed stadiometer and body weight (Tanita-BC 418 MA, Tanita Corporation, Tokyo, Japan).

### Smoking assessment

Smoking status was evaluated with an interviewer-administered questionnaire and collapsed into two categories for statistical analyses: Smokers (current and smokers during the last three months before pregnancy) vs non-smokers (former smokers and never smokers).

### DNA methylation profiling and genotyping

DNA was extracted consecutively throughout the data collection, at the Hormone Laboratory, Oslo University Hospital, using a salting out procedure [20]. The genome-wide DNA methylation measurement and genotyping were performed at the Department of Clinical Sciences, Clinical Research Centre, Lund University, Malmö, Sweden and have been described in detail elsewhere [15].

We quantified DNA methylation in peripheral blood leukocytes using the Infinium MethylationEPIC BeadChip (Illumina, San Diego, CA, USA). We used the Meffil R package [21] for Quality control (QC). 472 individuals from the 480 available, and 864,560 probes passed the QC. Blood cell composition (CD8T, CD4T, NK, Monocytes, B-cells and Neutrophils) were calculated with Meffil using Houseman’s reference panel [22] during the QC procedure. Lastly, Y and X chromosome probes that harbour SNPs and cross-reactive probes per Pidsley and collaborators’ list [23] were not considered for statistical analyses. Thus, the EWAS analyses performed are based on 792,530 probes in total.

Technical validation was performed on four CpG sites preliminary associated with fasting glucose, 2-hour glucose and BMI. Overall, the four sites had a good agreement between pyrosequencing and the EPIC array [15].

We used the Illumina CoreExome chip for genotyping and PLINK 1.9 software [24] for QC and variant filtering. Variants with deviations of Hardy Weinberg (HW) equilibrium (p=1.0×10^−6^), low call rate (<95%), and low minor allele frequencies (MAF), were removed. Around 300,000 variants were left for imputation and 300 European and 138 South Asian passed the QC. Imputation for Europeans and South Asians was done by mapping the GWAS scaffold to NBI build 37 of the human genome. For each ancestry we used their correspondent 1000 genomes project panel (Phase 3, - http://www.well.ox.ac.uk/~wrayner/tools/) [25], using IMPUTE2 (version 2.3.2) [26]. PLINK 1.9 was used for a post-imputation QC. We removed non-SNPs variants and low-quality post-imputation SNPs (info<0.9). We also filtered variants using stricter HW equilibrium (p-value<1.7×10^−07^) and MAF cutoffs (>5%).

We have previously assessed genetic ethnic origin through ancestry principal component analysis by using the variance-standardized relationship matrix using PLINK 1.9. There was a clear separation between EUR and SA which corresponded to their inferred ancestry [15]. We, therefore, use self-reported ancestry for the categorization of ethnicity.

### Study Flow

For the EWAS studies, we included 463 women (302 European and 161 South Asians) that passed the EWAS QC procedure and had serum folate available (Figure 1). From these, 295 samples from Europeans and 161 South Asians also passed the GWAS QC and could be included in the mQTL analysis (Figure 1).

**Figure 1:**
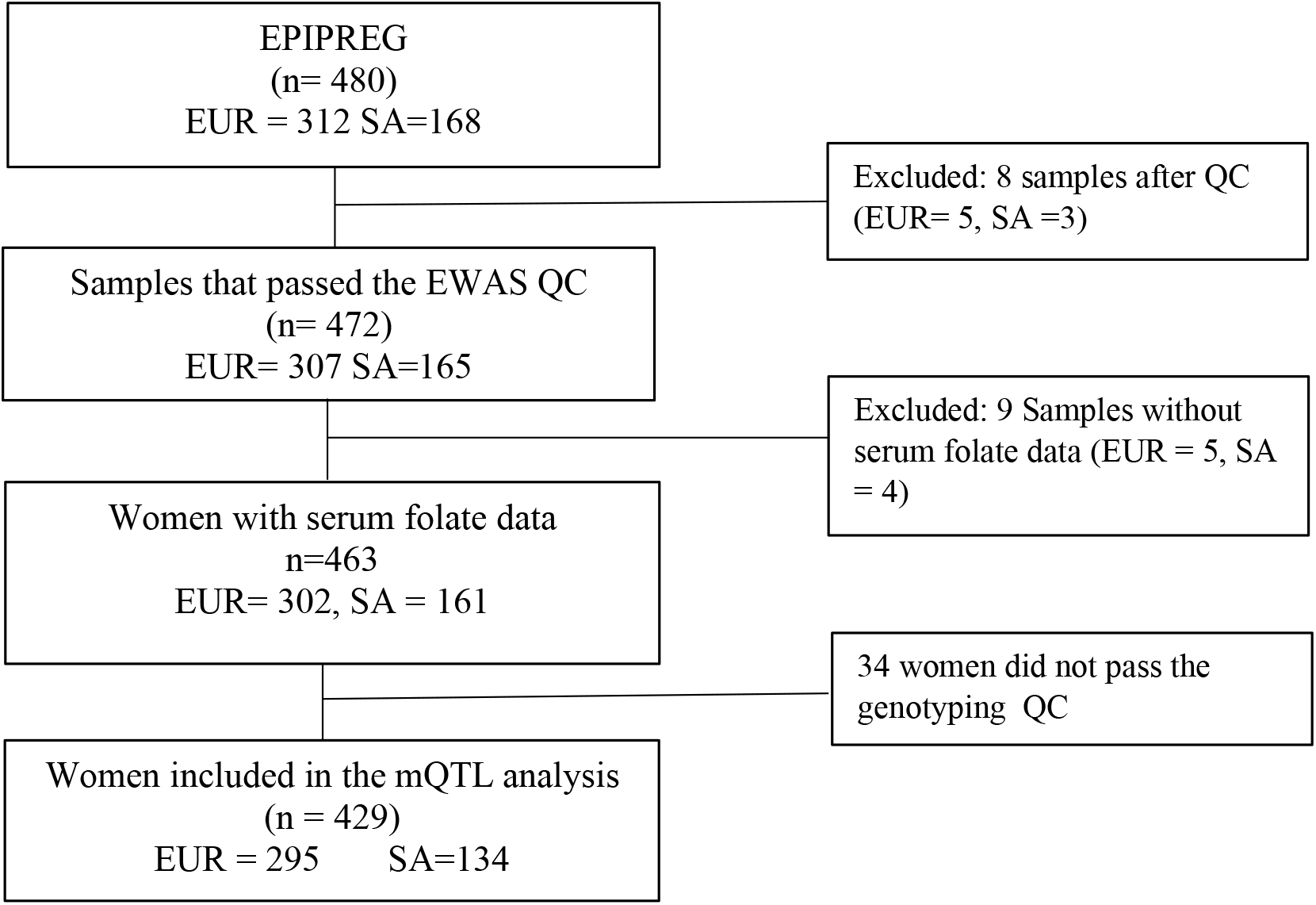
Study flow diagram that shows the samples selected for the EWAS analyses. EUR = Europeans, SA= South Asians.

### Statistics

Beta values were transformed to M-values [27]. For the cross-ancestry EWAS of serum folate levels, we used linear mixed models using the R packages lme4 and lmerTest [28], calculating the p-values with Satterthwaite’s method. The M-values were the outcome, and serum folate the exposure. The model was adjusted for Age, smoking and blood cell composition were used and ancestry was used as a random intercept. Ancestry-specific EWAS were performed separately in Europeans and South Asians using standard multivariate linear models using the *limma* R package [29], adjusting for age, smoking and blood cell composition. We use an FDR threshold of 5%.

For the Gene ontology (GO) analysis, we performed an over-representation test using the nominal significant CpG sites (p-value <1.0×10^−4^) with the web-software WebGestalt [30]. This was done for each EWAS analysis. We used “Biological process non-redundant” as the database of reference, and we only included GO terms that were composed of at least five genes. For this analysis, we accepted an FDR of 5%, but the 20 most significant GO terms with a p-value <0.05 are also reported.

For CpG hits from the cross-ancestry EWAS, we assessed their associations with cardiometabolic traits using linear mixed models and the same covariates as in the discovery analysis. For CpG hits from the ancestry-specific EWAS, we tested associations with cardiometabolic traits using standard linear regression models restricted to the same ancestry, including the same covariates from the corresponding EWAS. We accepted a p-value<0.05 for these analyses.

The GEM R package [31] was used for methyl quantitative trait loci (mQTL) analysis. For the cross-ancestry CpG sites, we performed the analysis separately in Europeans and South Asians, adjusting for age, smoking and blood cell composition, followed by a fixed effect meta-analysis. For ancestry-specific CpG sites, we analyzed the corresponding ancestry, using standard multivariate models adjusted for the abovementioned covariates. We used a standard genome-wide p-value threshold of 5×10^−8^. The mQTLs were defined as *cis*-mQTLs if they were located +-<1Mb from the methylation site, otherwise, they were classified as *trans*-mQTLs. The mQTLs were filtered by doing linkage disequilibrium analysis (LD)(R^2^>0.2) with the web-tool LD-link (https://ldlink.nci.nih.gov/?tab=home) [32]. As reference populations, we used Utah Residents from North and West Europe (CEU) for Europeans, and for South Asians Punjabi from Lahore, Pakistan (PJL) and Sri Lankan Tamil from the UK (STU). The mQTLs that survived the filtering analysis were tested for association (p-value<0.05) with cardiometabolic phenotypes in our sample. If the mQTL correspond to a cross-ancestry CpG site, we used mixed linear models with ancestry as random intercept for the SNP-phenotype analysis, otherwise we did the analysis in the corresponding ancestry where the CpG find was found by using simple linear models.

We used the R package mediation [33] for causal inference analyses to assess if DNA methylation mediated the relationship between an mQTL and serum folate, or if serum folate mediated the relationship between the mQTLs and DNA methylation. We included cell composition, age and smoking as covariates and 20,000 simulations were used to calculate the confidence intervals using a quasi-Bayesian approximation. A p-value <0.05 threshold was used for these analyses. The mediation analysis calculates the following models: The average causal mediation effects (ACME) which calculate the effect sizes when taking into account the mediator, and the average direct effect (ADE) which represents the effect size when adjusting for the mediator i.e. removing its effect, and the total effect which consists in the sum of ACME and ADE effect sizes.

### Replication in independent cohorts

We attempted replication of our results by using cross-sectional data of estimated folate intake calculated from a FFQ and DNA methylation of maternal peripheral blood leukocytes in two separate sub samples from The Norwegian Mother, Father and Child Cohort Study (MoBa) [34,35]. One sub-sample consisted of 1111 mothers, and 870 women assisted with reproductive technology (ART), and the other of 1022 mothers. Detailed methods and statistics used in MoBa can be consulted in the supplementary material.

We also attempted to replicate our findings using the results from an EWAS of maternal serum folate in cord-blood DNA methylation data, from a meta-analysis of a sub-sample of MoBa (N=1275) and Generation R (n=713) cohorts [12]. Details about the data analysis can be found in the source publication [12], but briefly, robust linear regression models were performed, adjusting for maternal age, maternal education, maternal sustained smoking during pregnancy, parity, and batch. We considered a CpG to be replicated if the CpG sites had consistent effects across tissues and a p<0.05.

We also evaluated if any of the 443 CpG (FDR<0.05%) sites reported by Joubert and collaborators [12], were replicated in EPIPREG data. We considered a CpG site as replicated if the CpG site had the same direction of the effect across both tissues and passed a Bonferroni p-value threshold (0.05/443). The CpG sites that reached a nominal p<0.05 are reported as suggestive findings.

### Consultations in public databases

We consulted the EWAS catalogue (http://ewascatalog.org/) [36] to look if the found CpG sites have been previously associated with folate or any cardiometabolic trait in peripheral blood leukocytes. The catalogue reports associations with a p<1×10^−4^. We used mQTLdb (http://www.mqtldb.org/) [37] to identify if the found CpG sites had mQTLs in maternal peripheral blood leukocytes. GoDMC database (http://www.godmc.org.uk/) [38], which consists of a meta-analysis of several cohorts with methylation at peripheral blood leukocytes, was also used to look for mQTLs as well. We report mQTLs that passed a standard p<5×10^−8^ threshold. We used PhenoScanner (http://www.phenoscanner.medschl.cam.ac.uk/) [39] to identify if the mQTLs identified in our sample or public databases were previously associated (p<5×10^−8^) with folate or any cardiometabolic phenotype.

## Results

### Population characteristics

Clinical characteristics of the women included are shown in Table 1.

**Table 1:**
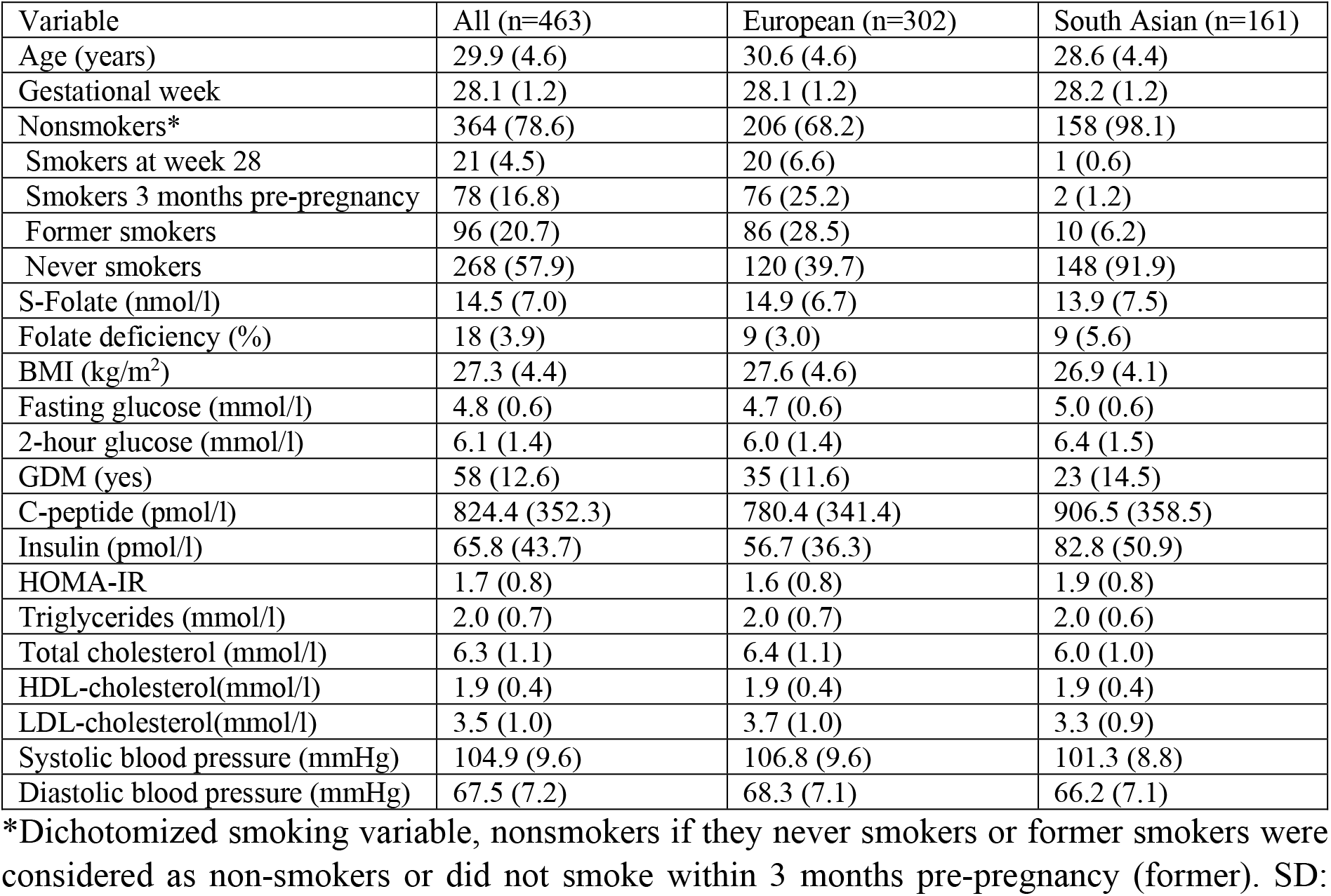
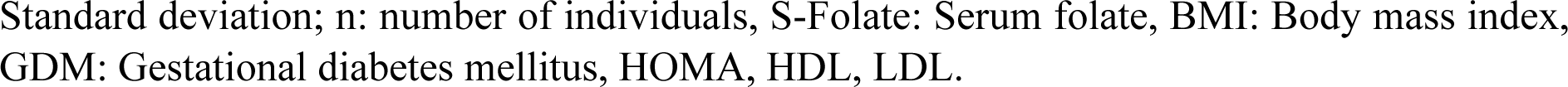
Clinical characteristics in gestational week 28 of the women included in the study. Data are presented as mean (SD) or n (%).

| Variable | All (n=463) | European (n=302) | South Asian (n=161) |
| --- | --- | --- | --- |
| Age (years) | 29.9 (4.6) | 30.6 (4.6) | 28.6 (4.4) |
| Gestational week | 28.1 (1.2) | 28.1 (1.2) | 28.2 (1.2) |
| Nonsmokers* | 364 (78.6) | 206 (68.2) | 158 (98.1) |
| Smokers at week 28 | 21 (4.5) | 20 (6.6) | 1 (0.6) |
| Smokers 3 months pre-pregnancy | 78 (16.8) | 76 (25.2) | 2 (1.2) |
| Former smokers | 96 (20.7) | 86 (28.5) | 10 (6.2) |
| Never smokers | 268 (57.9) | 120 (39.7) | 148 (91.9) |
| S-Folate (nmol/l) | 14.5 (7.0) | 14.9 (6.7) | 13.9 (7.5) |
| Folate deficiency (%) | 18 (3.9) | 9 (3.0) | 9 (5.6) |
| BMI (kg/m<sup>2</sup>) | 27.3 (4.4) | 27.6 (4.6) | 26.9 (4.1) |
| Fasting glucose (mmol/l) | 4.8 (0.6) | 4.7 (0.6) | 5.0 (0.6) |
| 2-hour glucose (mmol/l) | 6.1 (1.4) | 6.0 (1.4) | 6.4 (1.5) |
| GDM (yes) | 58 (12.6) | 35 (11.6) | 23 (14.5) |
| C-peptide (pmol/l) | 824.4 (352.3) | 780.4 (341.4) | 906.5 (358.5) |
| Insulin (pmol/l) | 65.8 (43.7) | 56.7 (36.3) | 82.8 (50.9) |
| HOMA-IR | 1.7 (0.8) | 1.6 (0.8) | 1.9 (0.8) |
| Triglycerides (mmol/l) | 2.0 (0.7) | 2.0 (0.7) | 2.0 (0.6) |
| Total cholesterol (mmol/l) | 6.3 (1.1) | 6.4 (1.1) | 6.0 (1.0) |
| HDL-cholesterol (mmol/l) | 1.9 (0.4) | 1.9 (0.4) | 1.9 (0.4) |
| LDL-cholesterol (mmol/l) | 3.5 (1.0) | 3.7 (1.0) | 3.3 (0.9) |
| Systolic blood pressure (mmHg) | 104.9 (9.6) | 106.8 (9.6) | 101.3 (8.8) |
| Diastolic blood pressure (mmHg) | 67.5 (7.2) | 68.3 (7.1) | 66.2 (7.1) |

\*Dichotomized smoking variable, nonsmokers if they never smokers or former smokers were considered as non-smokers or did not smoke within 3 months pre-pregnancy (former). SD:
Standard deviation; n: number of individuals, S-Folate: Serum folate, BMI: Body mass index, GDM: Gestational diabetes mellitus, HOMA, HDL, LDL.

### Discovery analysis in EPIPREG

In the cross-ancestry EWAS, increased methylation of cg19888088 (annotated to *EBF3*) and cg10871182 was associated with lower levels of serum folate (Figure 2 and Table 2). We did not find evidence of inflation (λ=1.05). Summary statistics of cross-ancestry CpG sites with p<1.0×10^−4^ are presented in Supplementary Table 1.

**Figure 2:**
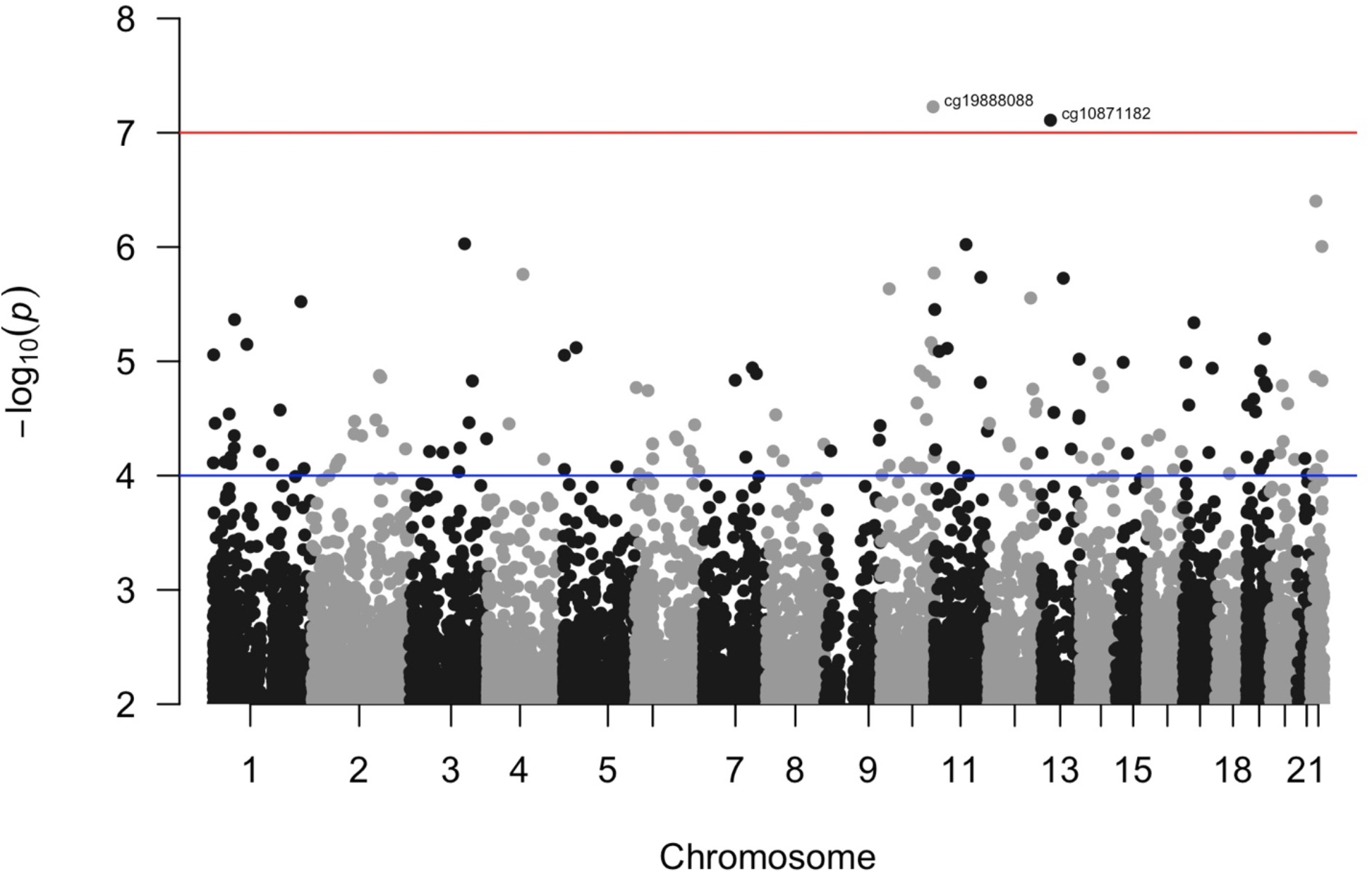
Manhattan plot of the cross-ancestry EWAS. Two CpG sites passed the FDR threshold (above the red line) of 5%. The blue line represents the nominal threshold (p-value<1.0×10^−4^).

**Table 2:** Summary statistics and genomic location of the CpG sites that passed the FDR<0.05 in each EWAS study.

| Cross-ancestry EWAS |  |  |  |  |  |  |  |
| --- | --- | --- | --- | --- | --- | --- | --- |
| CpG | Effect (SE) | p-value | FDR | UCSC Gene name | CHR:POS | Relation to Island | Relation to transcription |
| cg19888088 | -0.006 (0.001) | $5.94 \times 10^{-8}$ | 0.031 | *EBF3* | chr10:131686208 | North Shore | Body |
| cg10871182* | -0.019 (0.004) | $7.78 \times 10^{-8}$ | 0.031 | N/A | chr13:41959831 | Open Sea | N/A |
| European only EWAS |  |  |  |  |  |  |  |
| cg01952260 | -0.013 (0.002) | $3.88 \times 10^{-8}$ | 0.031 | N/A | chr9:138156618 | North Shore | N/A |
| South Asian only EWAS |  |  |  |  |  |  |  |
| cg07077240 | -0.024 (0.004) | $7.54 \times 10^{-8}$ | 6.0E-04 | *HERC3* | chr4:89514179 | Island | 5UTR |

\*CpG site driven by methylation outliers

In the ancestry specific EWAS, cg01952260 in Europeans (Table 2 and Supplementary Figure 1a) and cg07077240 (annotated in *HERC3*) in South Asians (Table 2 and Supplementary Figure 1b) were associated with reduced methylation at higher serum folate. We did not find evidence of inflation in either the European (λ=1.07) or South Asian EWAS (λ=1.06). Summary statistics of CpG sites with p<1×10^−4^ from the ancestry-specific EWAS are presented in Supplementary Table 2 (Europeans) and 3 (South Asians).,

Residual inspection of the models (Supplementary Figure 2) suggested that the association with cg10871182 was driven by outliers, thus it was not followed further.

### GO enrichment analysis

The GO-term “photoreceptor cell differentiation” passed the FDR<0.05 threshold (Figure 3 and Supplementary Table 4) when using the nominal probes from the cross-ancestry EWAS. The GO enrichment analysis in the European (Supplementary Table 5) EWAS did not show GO terms above the FDR threshold. Using the nominal CpG sites from the South Asian EWAS we found enrichment (FDR<0.05) in “cell-cell adhesion via plasma-membrane adhesion molecules” and “protein localization to cell periphery” (Supplementary Table 6).

**Figure 3:**
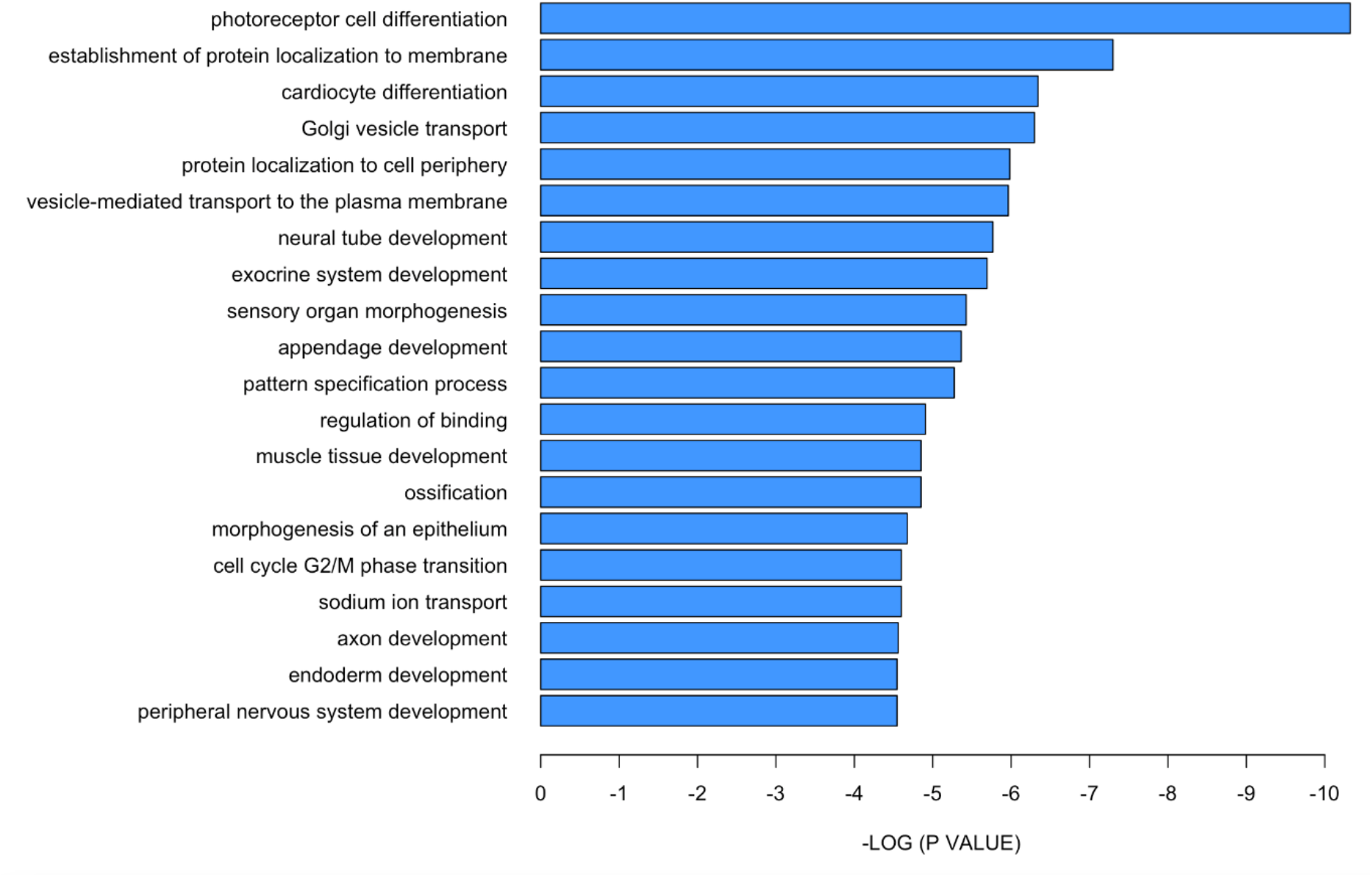
-log p-value bar plot of the 20 most significant GO-terms when using the nominal significant probes from the cross-ancestry EWAS. Only “photoreceptor cell differentiation” passed the FDR threshold of 5%, but the 20 most significant GO terms (uncorrected p<0.05) are also presented.

### Associations with cardiometabolic traits

cg19888088 identified in the cross-ancestry EWAS (effect size=-0.002, SE= 0.001, p-value = 0.0356), and cg01952260 identified in the European EWAS (effect size= 0.005, SE= 0.002, p-value = 0.022) were associated with diastolic blood pressure in EPIPREG (n=).Methylation levels of cg07077240 were not related to any of the other phenotypes tested (Supplementary Table 7).

According to the EWAS catalogue, cg19888088, cg01952260 and cg07077240 have not been reported to be associated with folate nor any cardiometabolic phenotype.

### Genetic variants and DNA methylation

In EPIPREG, mQTL analysis identified gene variants only for cg07077240 (rs12526070 and rs113796915) (Table 3). Because the variants were in strong LD with each other (R^2^=0.966) only rs12526070 was analyzed further for the phenotype associations. In EPIPREG, the minor allele of rs12526070 (T) was associated with increased serum levels of folate (p=9.19×10^−5^), triglycerides (p=0.006), total cholesterol (p=0.041), and HDL cholesterol (p=0.023) (Supplementary Table 8). We did not find associations between rs12526070 and serum folate levels nor with any cardiometabolic phenotypes in PhenoScanner.

**Table 3:** mQTLs associated with methylation at the folate CpG sites.

| CpG | mQTL | Gene | NE/E | CIS/TRANS | CHR:POS | EFFECT(SE) | p-value |
| --- | --- | --- | --- | --- | --- | --- | --- |
| cg07077240 | rs12526070 | *CASC15* | C/T | TRANS | 6:22272393 | -0.466 (0.079) | 3.67x10<sup>-8</sup> |
|  | rs113796915 | *CASC15* | A/G | TRANS | 6:22272564 | -0.466 (0.079) | 3.67x10<sup>-8</sup> |
| cg19888088 | N/A | N/A | N/A | N/A | N/A | N/A | N/A |
| cg01952260 | N/A | N/A | N/A | N/A | N/A | N/A | N/A |

\*NE: Non-effect allele, E: Effect allele, CHR: Chromosome, POS: Position

In mQTLdb, we found 4 trans-mQTL for cg07077240 (Supplementary Table 9) in LD within each other (R^2^ from 0.966 to 0.974). The most significant variant was rs4268751 locaed in *HS3ST4*. The variants rs112030070 and rs112571402 (R^2^=1) located in *WDR76* were trans-mQTLs for cg01952260 (Supplementary Table 9). GoDMC has not reported mQTLs for these CpG sites. PhenoScanner has not reported associations with folate nor any cardiometabolic phenotype for rs4268751 and rs112030070.

### Causal inference analysis

The average causal mediation effects (ACME) for cg07077240 were stronger than the average direct effects (ADE) (Table 4). The ADE was stronger than the ACME when using folate as a mediator between rs12526070 and cg07077240. Therefore, it is more likely that cg07077240 mediates the association between rs12526070 and serum folate levels.

**Table 4:**
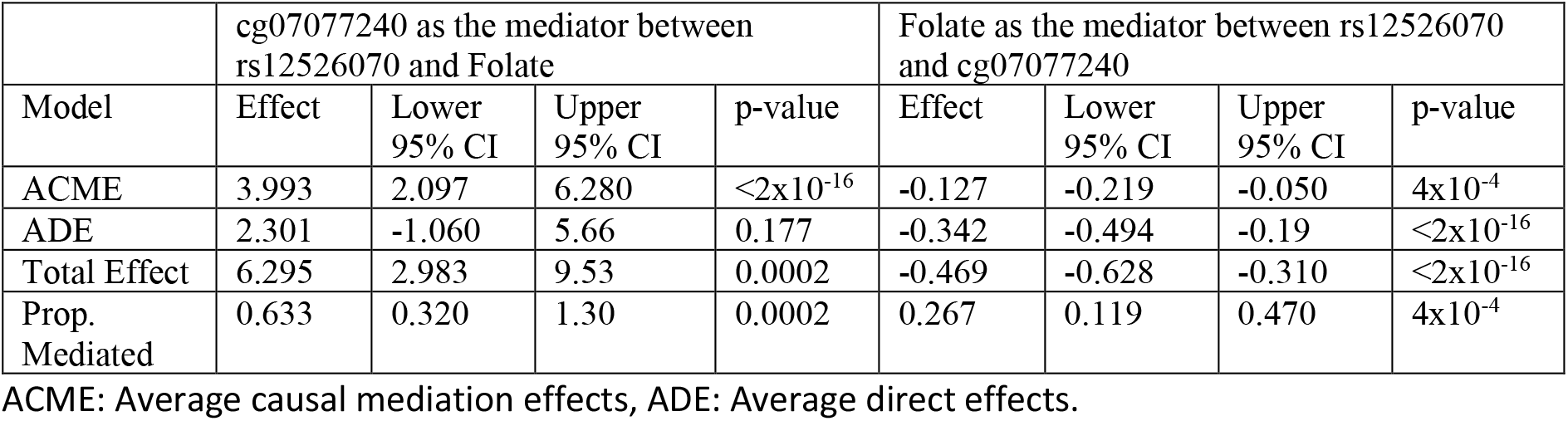
Mediation analysis to evaluate relationships between the rs12526070, methylation at cg07077240 and serum folate levels.

|  | cg07077240 as the mediator between rs12526070 and Folate |  |  |  | Folate as the mediator between rs12526070 and cg07077240 |  |  |  |
| --- | --- | --- | --- | --- | --- | --- | --- | --- |
| Model | Effect | Lower 95% CI | Upper 95% CI | p-value | Effect | Lower 95% CI | Upper 95% CI | p-value |
| ACME | 3.993 | 2.097 | 6.280 | <2x10<sup>-16</sup> | -0.127 | -0.219 | -0.050 | 4x10<sup>-4</sup> |
| ADE | 2.301 | -1.060 | 5.66 | 0.177 | -0.342 | -0.494 | -0.19 | <2x10<sup>-16</sup> |
| Total Effect | 6.295 | 2.983 | 9.53 | 0.0002 | -0.469 | -0.628 | -0.310 | <2x10<sup>-16</sup> |
| Prop. Mediated | 0.633 | 0.320 | 1.30 | 0.0002 | 0.267 | 0.119 | 0.470 | 4x10<sup>-4</sup> |

ACME: Average causal mediation effects, ADE: Average direct effects.

### Replication with folate intake data from MoBa

We did not find evidence of replication in any of the three CpG tested (Supplementary Table 10).

### Shared signatures between cord blood and maternal blood DNA methylation

None of the three CpG sites identified in EPIPREG in maternal blood were replicated in cord blood (Supplementary Table 10). Further, none of the CpG sites associated with serum folate in cord blood reached the Bonferroni threshold in EPIPREG, but 22 CpG sites had consistent effects across both tissues and a nominal p<0.05 in the cross-ancestry analysis (Supplementary Table 11). In the European and South Asian analyses, we found in addition 3 and 16 CpG sites that had consistent effects and a nominal p<0.05, respectively (Supplementary Tables 12 and 13).

## Discussion

We found that methylation in maternal peripheral blood leukocytes at cg19888088 (in *EBF3*), cg01952260 and cg07077240 (in *HERC3*) were inversely associated with maternal serum folate. Methylation at cg19888088 and cg01952260 were associated with diastolic blood pressure. rs12526070, mQTL of cg07077240 annotated to *CASC15* was associated with serum folate, triglycerides, total cholesterol, and HDL cholesterol. Mediation analysis suggested that cg07077240 mediates the relationship between rs12526070 and folate levels. Finally, we did no replicate neither three CpG sites in independent samples.

As no cohort had both serum folate and DNA methylation in the same individuals, we attempted replication in two samples with estimated folate intake from FFQ and DNA methylation in the same women, and in an EWAS of maternal serum folate and DNA methylation in cord blood of their offspring. Nevertheless, we did not replicate our findings. Despite serum folate and folate intake evaluated with FFQ are modestly correlated [40-42], FFQ could be prone to inaccurate reporting and other method limitations [43]. Therefore, the lack of replication could be due to underlying methodological differences across folate measurement methods. The lack of replication in cord blood could be because leukocyte composition between peripheral blood and cord blood differs [44], which may affect the associations. Furthermore, serum folate was not measured in the same individual i.e., the offspring but in maternal blood serum. The transport of maternal folate to cord blood depends on the activity of several transporters in the placenta [45]. During gestation, folate is accumulated in the placenta and cord blood to ensure enough folate to the fetus [45,46], thus folate levels in the fetus can be higher than maternal folate levels. Hence, in this context, maternal folate levels are an indirect measurement of folate in cord blood, which may affect our replication efforts. Studies with serum folate and DNA methylation data in peripheral blood leukocytes, and larger study populations are necessary to further confirm our findings.

The cross-ancestry site cg19888088, located in the *EBF3* gene, has not previously been associated with serum folate levels. However, DNA methylation at two other CpG sites within *EBF3* (cg26229752 and cg17726092) in cord blood leukocytes have been inversely associated with maternal serum folate levels [12]. Thus, we speculate that maternal folate levels are associated with lower *EBF3* methylation levels. *EBF3* is a transcription factor that influences the cerebral cortex’s laminal formation [47]. Loss of function mutations in *EBF3* have been associated with developmental defects such as altered neural development [48], and phenotypes such as intellectual disability, facial dysmorphism, ataxia, and autism [47,49]. Evidence further suggests that *EBF3* may act as a tumor suppressor [50]. Both aberrant DNA methylation patterns [50,51] and gene silencing in *EBF3* [50] are associated with several types of cancers.

The site cg01952260 found in Europeans and cg07077240 found in South Asians have not previously been associated with folate. Decreased DNA methylation at cg01952260 in naïve CD4+ T cells has been associated with systemic lupus erythematosus [52]. cg07077240 is located in the *HERC3* gene, that negatively regulates the NF-Kb pathway which is an important activator of inflammatory and immune reactions [53]. Folate is a vital nutrient for regulation of the immune reaction and inflammatory response [54]. Therefore, methylation at these CpG sites could potentially play a role in inflammatory related processes associated with folate.

Methylation levels of cg01952260 and cg07077240 were associated with diastolic blood pressure in EPIPREG. Folate has previously been associated with blood pressure [55,56]. Furthermore, a meta-analysis found that intake of folic acid in pregnancy lowered preeclampsia risk, more specifically through multivitamins containing folic acid [57] or other folic acid supplements [58]. Nevertheless, only Yadav and collaborators [59] has suggested that DNA methylation could be an intermediate mechanism between folate metabolism and blood pressure. The study found that global hypomethylation was associated with blood pressure in a North Indian population. The association was more pronounced in non-medicated hypertensive individuals carrying the T allele of the variant rs1801133 located in *MTHFR* [59]. Interestingly, another study found that BMI was associated with DNA methylation in the *MTHFS* gene, an enzyme relevant to folate metabolism [60]. Hence, folate regulation could be associated with DNA methylation patterns that in turn were associated with other cardiometabolic related traits. However, in the EWAS catalogue, we did not find previous associations between blood pressure and methylation at cg01952260 and cg07077240. Hence, these finding should be corroborated in other studies.

rs12526070, mQTL of cg07077240, was associated with several lipid parameters, which have not been reported previously. However, in pregnancy, the lipid profile fluctuates, with a decrease in the first trimester followed by a subsequent increase as pregnancy progress [61]. Therefore, it remains to be seen if the associations between rs12526070 and lipids are related to the lipid traits themselves or represent pregnancy-related lipid changes. The mQTLs previously reported for cg07077240 and cg01952260 in mQTLdb were not replicated in our mQTL analysis, nor in the GoDMC database. Further studies are needed to verify the mQTLs associated with cg07077240 and cg01952260.

The mediation analysis implied that the T allele of rs12526070 increases folate levels, through methylation at cg07077240 and probably by altering the gene expression of *HERC3*. Nevertheless, other unknown environmental factors may alter cg07077240 methylation as well, which could regulate folate levels independently of the rs12526070. Unfortunately, we lack independent variants to perform a robust Mendelian randomization analysis to establish causal relationships.

A major strength of this study is the well characterized cohort which allowed association analyses with other cardiometabolic phenotypes. Further, genetic data allowed us to perform mQTL analysis to find potential gene variants related to DNA methylation of the CpG sites. An important limitation of this work is the small sample size, only allowing detection of moderate to strong effect sizes. Finally, we lacked a proper replication cohort with DNA methylation in peripheral blood leukocytes and serum folate data in the same individuals.

## Conclusion

We identified three novel CpG sites associated with serum folate levels in peripheral blood leukocytes. The cross-ancestry CpG site cg19888088 was located in the *EBF3* gene, a gene related to neural development. cg01952260, found in Europeans, and cg07077240 (in *HERC3*) found in South Asians, are potentially associated with immune processes related to inflammation. Methylation at cg19888088 and cg01952260 was associated with diastolic blood pressure. Only cg07077240’s was associated with genetic variants, and its mQTL, rs12526070 (in *CASC15*), was associated with serum folate and lipids. In overall, our findings provide new insights about the epigenomic component of serum folate levels.

## Future perspective

Our findings should be replicated in larger cohorts with both serum folate and DNA methylation data in peripheral blood leukocytes.

## Summary points

- We identified three CpG sites associated with serum folate in peripheral blood leukocytes: cg19888088 (cross-ancestry), cg01952260 (in Europeans), and cg07077240 (in South Asians).
- cg19888088 is located in the *EBF3* gene, which has been implicated in neural development, and methylation at *EBF3* has been previously associated with maternal serum folate levels in cord blood.
- cg01952260 and cg07077240 in *HERC3* are potentially associated with immune processes related to inflammation.
- cg19888088 and cg01952260 were associated with diastolic blood pressure in EPIPREG.
- cg07077240’s mQTL, rs12526070 (annotated in *CASC15*), was associated with serum folate and lipids.
- A proper replication cohort with both serum folate and DNA methylation data in peripheral blood leukocytes is needed to verify our findings.

## Data Availability

Due to strict regulations for genetic data and privacy protection of patients in Norway, all requests for data access are processed by the STORK Groruddalen project's steering committee. Please contact the PI of STORK Groruddalen or the PI of EPIPREG.
Data from the Norwegian Mother, Father and Child Cohort Study and the Medical Birth Registry of Norway used in this study are managed by the national health register holders in Norway (Norwegian Institute of public health) and can be made available to researchers, provided approval from the Regional Committees for Medical and Health Research Ethics (REC), compliance with the EU General Data Protection Regulation (GDPR) and approval from the data owners. The consent given by the participants does not open for storage of data on an individual level in repositories or journals. Researchers who want access to data sets for replication should apply through helsedata.no. Access to data sets requires approval from The Regional Committee for Medical and Health Research Ethics in Norway and an agreement with MoBa.

## Author contributions

NFB and CS contributed to the study conceptualization and design of this sub-study. CS and KIB conceptualized and designed the EPIPREG sample. NFB conducted the statistical analyses in EPIPREG, drafted the manuscript, and performed the post-imputation QC. CMP performed the replication analysis in MoBa. BJ and SL provided the summary statistics for the replication analysis in cord blood. SLØ curated EPIPREG data. RBP facilitated the wet lab experiments regarding the methylation and genotyping chips. GHM performed the QC of the genomic data and imputation in EPIPREG. KIB and AKJ designed the STORK-G project. LS contributed with data acquisition in STORK-G. CS is the guarantor of this work, had access to the data and accepts full responsibility for the conduct of the study. All coauthors reviewed/edited the manuscript and approved the final version.

## Acknowledgements

We would like to thank the women who participated in the STORK Groruddalen study, Maria Sterner, Malin Neptin, and Gabriella Gremsperger at the Genomics Diabetes and Endocrinology CRC, Malmö, for the wet lab experiments of the bead chips, and Leif C. Groop, Lund University Diabetes Centre, Malmö, Sweden, for facilitating the wet lab experiments.

## Data availability

Due to strict regulations for genetic data and privacy protection of patients in Norway, all requests for data access are processed by the STORK Groruddalen project’s steering committee. Please contact the PI of STORK Groruddalen or the PI of EPIPREG.

Data from the Norwegian Mother, Father and Child Cohort Study and the Medical Birth Registry of Norway used in this study are managed by the national health register holders in Norway (Norwegian Institute of public health) and can be made available to researchers, provided approval from the Regional Committees for Medical and Health Research Ethics (REC), compliance with the EU General Data Protection Regulation (GDPR) and approval from the data owners. The consent given by the participants does not open for storage of data on an individual level in repositories or journals. Researchers who want access to data sets for replication should apply through helsedata.no. Access to data sets requires approval from The Regional Committee for Medical and Health Research Ethics in Norway and an agreement with MoBa.

