## supplementary material for "Epigenome-wide association study of serum folate in maternal peripheral blood leukocytes"

### Supplementary methods

#### The Norwegian Mother, Father and Child Cohort Study (MoBa)

MoBa is a cohort study which includes more than 114,000 children, 95,000 mothers and 75, 000 fathers [1]. Between 1999 and 2008 women were enrolled in the study during pregnancy, of which 40% of those invited participated. The participants are constantly followed with different questionnaires and by linkage to data associated with their Medical Birth Registry of Norway (MBRN) [2]. Blood samples from the parents were taken during pregnancy, and from the umbilical cord at delivery [1,3]. For this study, we selected two sets of women. Among the samples with both DNA methylation and folate intake available, one set consisted of 1000 mothers randomly selected, and 1000 matching assisted reproductive technology (ART) cases, and the other contained 1200 randomly selected mothers. Samples overlaping between the two sets were removed. The final sample size was 1981 in one sample, of which 870 mothers received ART treatment, and 1022 randomly selected samples in the second set. MoBa's methods regarding how ART was defined, DNA methylation measurement, and folate estimation with FFQ can be consulted in the supplementary material.

#### Subfertility and ART

It is mandatory for fertility clinics to report ART procedures to the birth registry, such as in vitro fertilization (IVF) with or without intracytoplasmic sperm injection (ISCI), and fresh or frozen thawed embryo transference. ART was defined as any ART except inseminations, and coded as dichotomic variable [1].

#### DNA methylation processing

DNA samples were delivered to the Institute of Life and Brain Sciences, University of Bonn, in Germany, where they were processed on the Illumina MethylationEPIC BeadChip array. The EZ-96DNA methylation-Lgihtning™ MagPrep kit (Zymo Reasearch, Irvine, USA), was used for bisulfite conversion of the DNA. Raw data from the MethylationEPIC BeadChip array was extracted with Genome Studio 2011.2. The iDAT files were processed for quality control with RnBeads R package in four separated batches. 44,210 cross-hybridizing probes, and 16,117 probes of which the last three bases overlapped with a SNP, were removed. Probes with a detection p-value >0.01 were also removed. Greedycut algorithm was used to evaluate potential batches, and further remove samples and probes with outlying DNA methylation. The rest of the DNA methylation signals were further corrected for background noise using the NOOB method . Signal intensity of the samples was visually inspected using the output of control probes obtained from RnBeads. CpG sites which were problematic per the batches presented in RnBeads's report were removed as well. In total 770,586 probes passed the QC and were considered for further analyses. BMIQ (Beta-mixture quantile normalization) method was used to normalize type I and type II probes.

**Measures of Folate intake and folic acid use**

MoBa food frequency questionnaire (FFQ) (available at: http://www.fhi.no/dav/011fbd699d.pdf) was mailed to all participants around gestational week 15TH, and completed around weeks 16-18th weeks of gestation [2]. It consist on a semi-quantitative questionnaire which asks about 255 food items and its designs allows to capture dietary habits and intake of dietary supplements during the first month of pregnancy. Details of the questionnaire has been described in detailed previously [2]. For the total folate intake (expressed as folic acid equivalents) both food folate and folic acid from supplements were considered [3].

**Statistical analyses**

For the first set, we fitted linear mixed models of the three discovered CpG sites using age, smoking and blood cell composition as fixed effects and whether the women went through in ART as a random intercept. For the second set, we performed standard multivariate linear models adjusted for age, smoking and cell composition. The analyses were done using the package *Rfast* [4]. A CpG was defined as replicated if it followed the same direction of the effect and had $p<0.05$.

**Supplementary Figures**

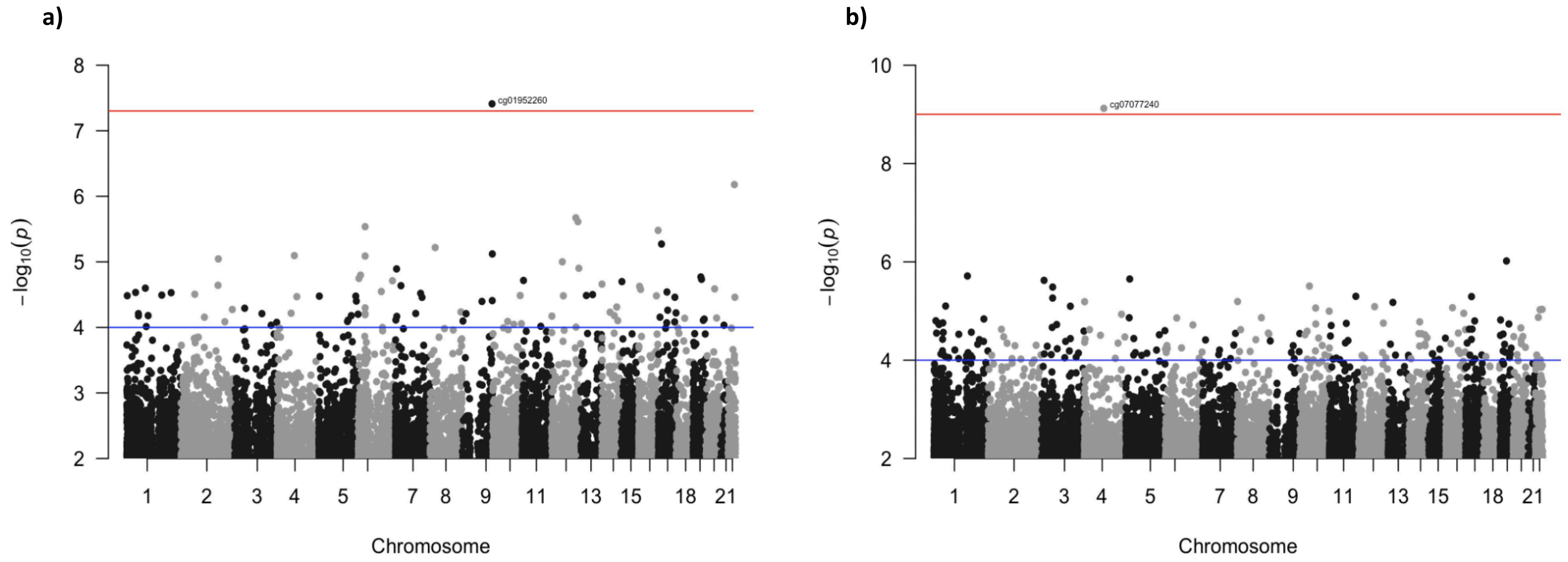


**Supplementary figure 1**: Manhattan plots showing the EWASes for the Europeans (a) and South Asians (b) separately. The CpG sites above the red line passed the FDR threshold of 5%. The blue line represent the nominal threshold (p-value<$1.0\times10^{-4}$).

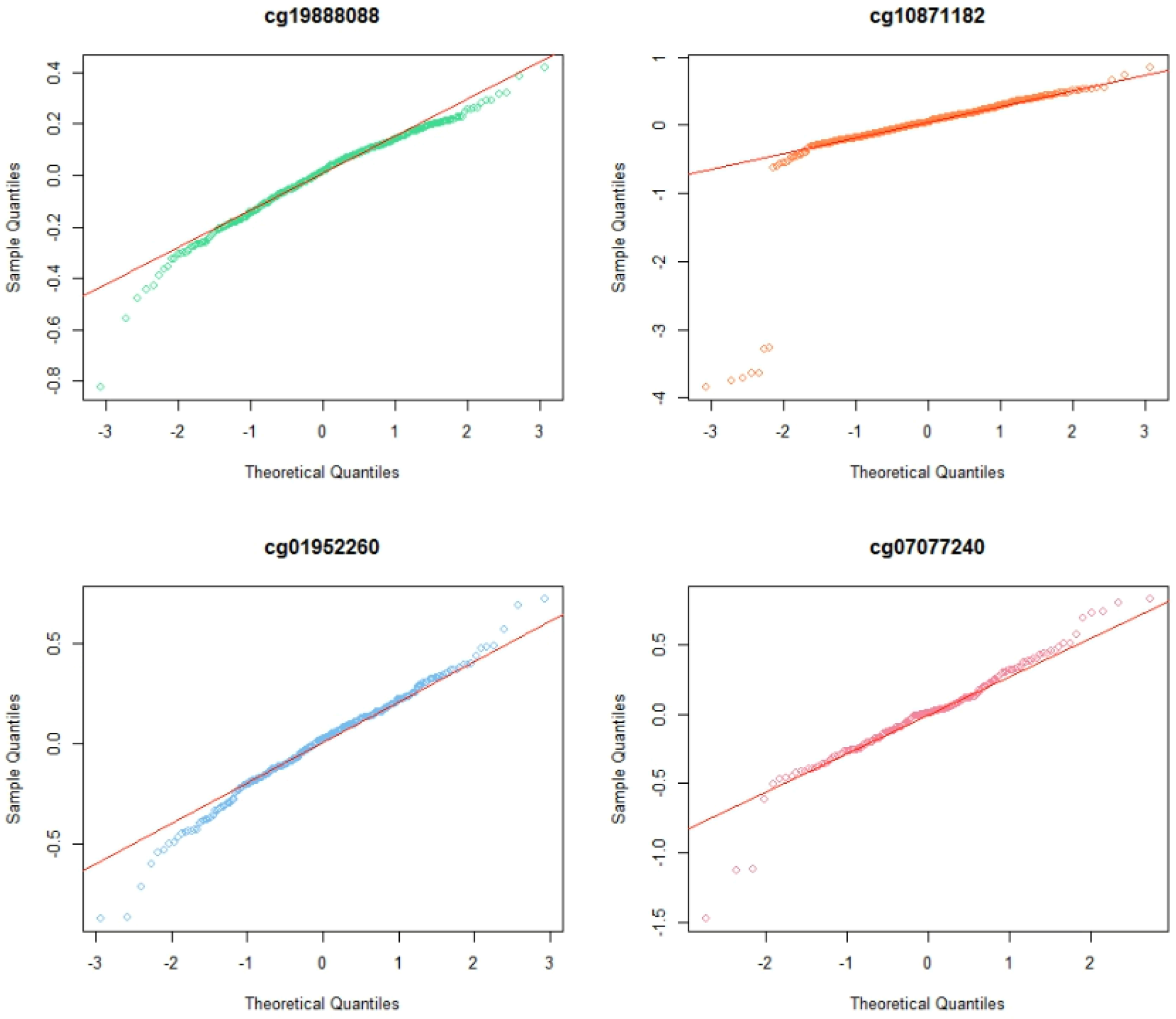


**Suplementary figure 2:** qqplots for the model´s residual for each CpG found in the EWAS analyses. It is noticiable that **cg10871182** have several extreme outliers in methylation.
